# Image-to-image generative adversarial networks for synthesizing perfusion parameter maps from DSC-MR images in cerebrovascular disease

**DOI:** 10.1101/2022.05.24.22274901

**Authors:** Tabea Kossen, Vince I Madai, Matthias A Mutke, Anja Hennemuth, Kristian Hildebrand, Jonas Behland, Adam Hilbert, Jan Sobesky, Martin Bendszus, Dietmar Frey

## Abstract

Stroke is a major cause for death or disability. As imaging based patient stratification improves acute stroke therapy, dynamic susceptibility contrast magnetic resonance imaging (DSC-MRI) is is of major interest to image brain perfusion. However, expert-level perfusion maps require a manual or semi-manual post-processing by a medical expert making the procedure time-consuming and less standardized. Modern machine learning methods such as generative adversarial networks (GANs) have the potential to automate the perfusion map generation on an expert-level without manual validation. We propose a modified pix2pix GAN with a temporal component (temp-pix2pix-GAN) that generates perfusion maps in an end-to-end fashion. We train our model on perfusion maps infused with expert knowledge to encode it into the GANs. The performance was trained and evaluated using the structural similarity index measure (SSIM) on two datasets including acute stroke patients and patients with steno-occlusive disease. Our temp-pix2pix architecture showed high performance on the acute stroke dataset for all perfusion maps (mean SSIM 0.92-0.99) and good performance on data including patients with steno-occlusive disease (mean SSIM 0.84-0.99). While clinical validation is still necessary in future studies, our results mark an important step towards automated expert-level perfusion maps and thus, fast patient stratification.

## 1 INTRODUCTION

Ischemic stroke is a leading cause for death or disability worldwide^1^. Standard treatment strategies include recanalization by mechanical or pharmacological intervention, or a combination of both (Berge et al. (2021); Turc et al. (2019)). In this context, the eligibility of patients for treatment is mainly based on large cohorts of interventional trials that implement few imaging information (Lin et al. (2022); McDermott et al. (2019)). However, this means that some patients will not receive treatment that would be of benefit for them and, conversely, some patients will be subjected to futile treatment attempts (Goyal et al. (2016)). An alternative approach to improve outcomes is an individualized patient stratification based on specific patient characteristics (Rehani et al. (2020); Sharobeam and Yan (2022)).

One of the most important techniques for this approach is perfusion weighted-imaging, a special imaging technique used in both computed tomography (CT) and magnetic resonance imaging (MRI) (Sharobeam and Yan (2022)). It provides highly relevant information about (patho)physiological blood flow in and around the ischemic brain tissue (Copen et al. (2011)). In MRI, the most commonly used perfusion imaging technique is dynamic susceptibility contrast (DSC) MRI (Jahng et al. (2014)). It measures brain perfusion by injecting a gadolinium-based contrast agent into the patient’s blood (Jahng et al. (2014)), followed by a series of T2- or T2*-weighted MRI sequences that record the flow of the contrast agent through the brain. The resulting 4D image is deconvolved voxel-wise with an arterial input function (AIF) (Calamante (2013)). The tissue concentration curve as well as the deconvolved curve result in interpretable perfusion parameter maps such as the cerebral blood flow (CBF), cerebral blood volume (CBV), mean transit time (MTT), time-to-maximum (Tmax), and time-to-peak (TTP) (Calamante (2013)). Importantly, the placement of the AIF is performed either in a semi-manual or manual manner to achieve the highest quality. Additionally, automated methods exist that in some areas -such as in stroke - require little input by experts to provide perfusion parameter maps of high quality (Hansen et al. (2016); Ben Alaya et al. (2022); Krusche et al. (2021)). In clinical practice, however, all existing methods of AIF determination require at least some oversight by experts to rule out faulty calculations due to suboptimal AIFs. This is a particular challenge in stroke care, where time is a critical resource as it is one of the most important determinants of clinical outcome. Therefore, there is a great clinical need for novel automation approaches that provide expert-level perfusion maps without the necessity for any manual input.

One possible solution is the application of modern artificial intelligence (AI) methods based on machine learning and here particularly deep learning approaches. These have shown great promise for solving medical imaging problems in the past years (Wernick et al. (2010); Lundervold and Lundervold (2019)). Among deep learning applications, generative adversarial networks (GANs) are particularly promising for the generation of expert level perfusion maps. For example, GANs can be presented both with an original image and a processed image and learn to generate the processed image from the original. This is achieved by the special architecture of GANs: They consist of two neural networks that try to fool each other (Goodfellow et al. (2014)). One network, the generator, synthesizes a data sample such as an image, whereas the other network, the discriminator, decides whether the sample looks like a real sample or not. At the end of the training, the generated sample should resemble the original as closely as possible. For image-to-image translations GANs are considered to be state-of-the-art in the medical field (Yi et al. (2019); Zhu et al. (2020)) and a conditional GAN such as the pix2pix GAN can be applied (Isola et al. (2018)). For example, pix2pix GANs have been successfully applied to transform MR images to CT images (cross-modal) or to transform 3T MR images to 7T MR images (intramodal) (Brou Boni et al. (2020); Nie et al. (2018)).

Given that the translation of a time-series of perfusion information from source images to a single perfusion map can be seen as a highly similar medical image-to-image translation problem, GANs are a highly promising method for this use case. Preliminary work on GANs for the translation of time-series in dynamic cine applications has been published (Ghodrati et al. (2021)). Yet, to the best of our knowledge no study has investigated the generation of DSC perfusion images from perfusion source data so far.

Thus, we propose a modified slice-wise pix2pix GAN with a temporal component (temp-pix2pix-GAN) to account for the time dimension in DSC source perfusion imaging. Our GAN model automatically generates perfusion parameter maps in an end-to-end fashion. We train our model on expert-level perfusion parameter maps (see Figure 1). The performance of our temp-pix2pix GAN model is compared to a standard pix2pix GAN without a temporal component. We train and test our approach on two different datasets including acute stroke patients as well as patients with chronic cerebrovascular disease.

**Figure 1.**
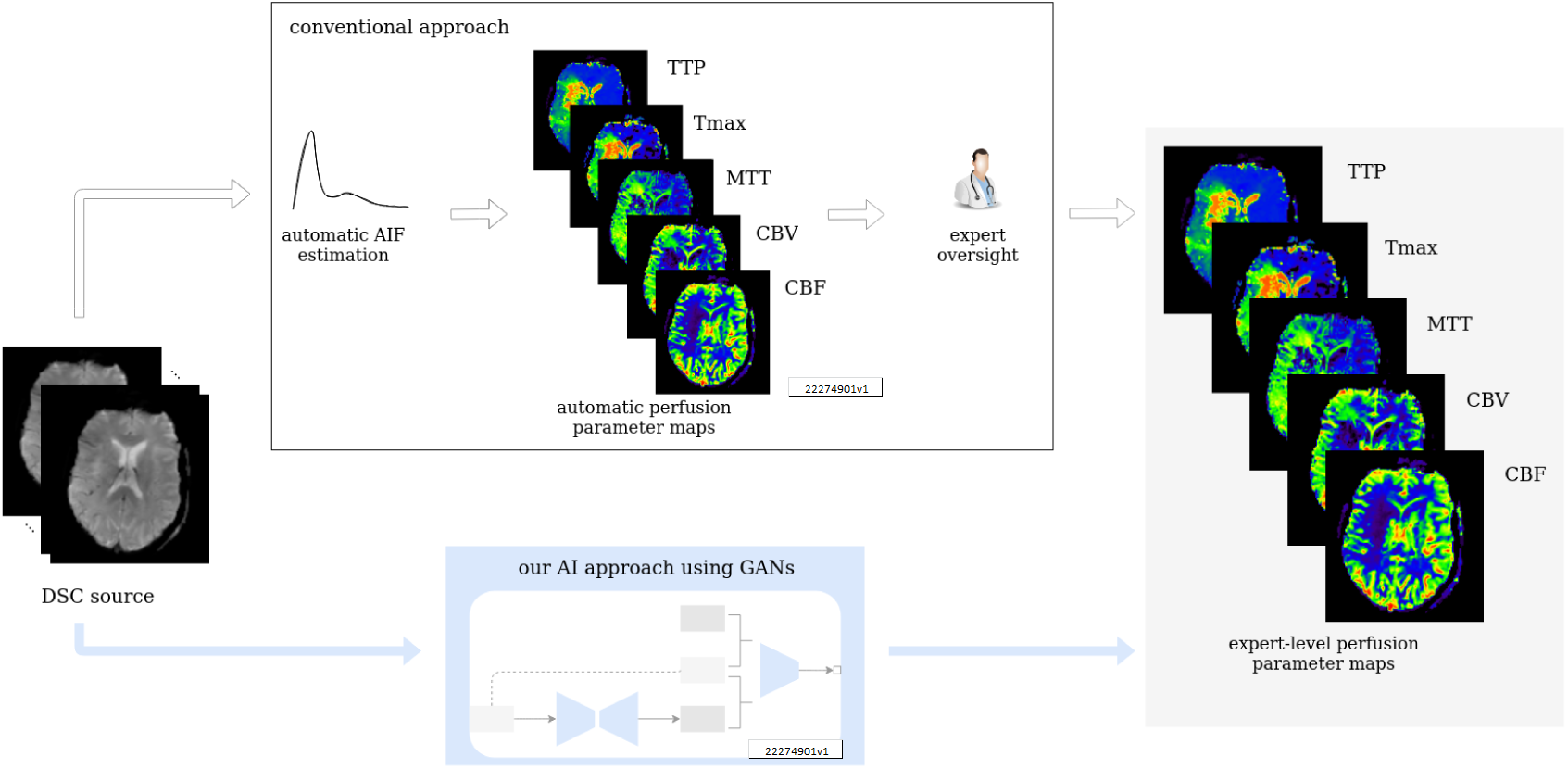
Workflow of study. Our GAN is trained on expert-level perfusion maps. The resulting model is able to synthesize perfusion maps from unseen data without the need of manual AIF selection, at the same expert level that was present in the training data.

## 2 MATERIALS AND METHODS

### 2.1 Data

In total, 276 patients were included in this study. 204 patients from study Heidelberg suffered from acute stroke. 204 patients from a study performed at Heidelberg University Hospital that suffered from acute stroke. Imaging was performed with a T2*-weighted gradient-echo EPI sequence with fat supression TR=2220ms, TE=36ms, flip angle 90°, field of view: 240×240mm^2^, image matrix: 128×128mm, 25-27 slices with ST of 5mm and was started simultaneously with bolus injection of a standard dose (0.1mmol/kg) of an intravenous gadolinium-based contrast agent on 3 Tesla MRI systems (Magnetom Verio, TIM Trio and Magnetom Prisma; Siemens Healthcare, Erlangen, Germany). In total, 50 to 75 dynamic measurements were performed (including at least eight prebolus measurements). Bolus and prebolus were injected with a pneumatically driven injection pump at an injection rate of 5ml/s. The study protocol for this retrospective analysis of our prospectively established stroke database was approved by the ethics committee of Heidelberg University and patient informed consent was waived.

72 patients with steno-occlusive disease were included from the PEGASUS study (Mutke et al. (2014)). 80 whole-brain images were recorded using a single-shot FID-EPI sequence (TR=1390ms, TE=29ms, voxel size: 1.8×1.8×5mm^3^) after injection of 5ml Gadovist (Gadobutrol, 1 M, Bayer Schering Pharma AG, Berlin) followed by 25ml saline flush by a power injector (Spectris, Medrad Inc., Warrendale PA, USA) at a rate of 5ml/s. The acquisition time was 1:54 minutes. All patients gave their written informed consent and the study has been authorized by the ethical review committee of Charite - Universitatsmedizin Berlin.

DSC post-processing was performed blinded to clinical outcome. For the acute stroke data from Heidelberg, DSC data were post-processed with Olea Sphere^®^ (Olea Medical, La Ciotat, France), automatic motion correction was applied. Raw DSC images were used to calculate perfusion maps of time-to-peak (TTP) from the tissue response curve. Maps of cerebral blood flow (CBF), cerebral blood volume (CBV), mean transit time (MTT), and time-to-maximum (Tmax) were created by deconvolution of a regional concentration time curve with an arterial input function (AIF). Block-circulant singular value decomposition (cSVD) deconvolution was applied. The arterial input function (AIF) was detected automatically. All AIFs were visually inspected by a neuroradiology expert (MAM, over 6 years experience in perfusion imaging) and only in two cases the automatically detected AIF needed to be manually corrected.

For PEGASUS patients, DSC data were post-processed with the PGui software (Version 1.0, provided for research purposes by the Center for functional neuroimaging, Aarhus University, Denmark). Motion correction was not available. Raw DSC images were used to calculate perfusion maps of TTP from the tissue response curve. Maps of CBF, CBV, MTT, and Tmax were created by deconvolution of a regional concentration time curve with an AIF. Parametric deconvolution was applied (Mouridsen et al. (2014)). For each patient, an AIF was determined by a junior rater (JB, 2 years experience in perfusion imaging) by manual selection of three or four intravascular voxels of the MCA M2 segment contralateral to the side of stenosis minimizing partial volume effects and bolus delay. The AIF shape was visually assessed for peak sharpness, bolus peak time and amplitude width (Calamante (2013); Thijs et al. (2004)). The AIFs were inspected by a senior rater (VIM, over 12 years of experience in perfusion imaging).

The post-processed data was split into a training (acute stroke data: 142, PEGASUS: 50 patients), validation (acute stroke data: 20, PEGASUS: 8 patients) and test (acute stroke data: 41, PEGASUS: 12 patients) set. The models were trained on the respective training set and the hyperparameters were selected based on the performance on the validation test. The generalizable performance was estimated by the performance of the test set. The acute stroke data was resized to 21 slices each containing 128×128 voxels. The DSC source was rescaled to 80 time points. All images of one parameter map as well as the DSC source images were normalized between -1 and +1 and split into slices.

### 2.2 General methodological approach

We utilized a special type of AI model that was developed for generating an image based on the input of another image: the pix2pix GAN (Isola et al. (2018)). A pix2pix GAN consists of two neural networks that try to mislead each other. The first network, the generator, aims to produce realistic looking images based on another image (e.g. produce a CT based on a MR image), whereas the second network, the discriminator, tries to distinguish between the generated and real images. Based on the discriminator’s feedback, both networks get better in their respective tasks.

Typically, the input and output to a pix2pix GAN generator is a 2D image. For this use-case, we modified the pix2pix GAN to take a 3D image (time sequence of the 2D DSC source image) as an input and synthesize the corresponding 2D perfusion map slice (e.g. Tmax). In this work we implemented two different generator architectures. The first architecture, the classical pix2pix GAN, took in the 3D input image without accounting for the temporal relation between the images. In contrast to that, the second architecture, the temp-pix2pix GAN, was designed to first extract the temporal relation between the images followed by the transformation to the output image (see Figure 2). In the following, the technical details of the two approaches are described in depth.

**Figure 2.**
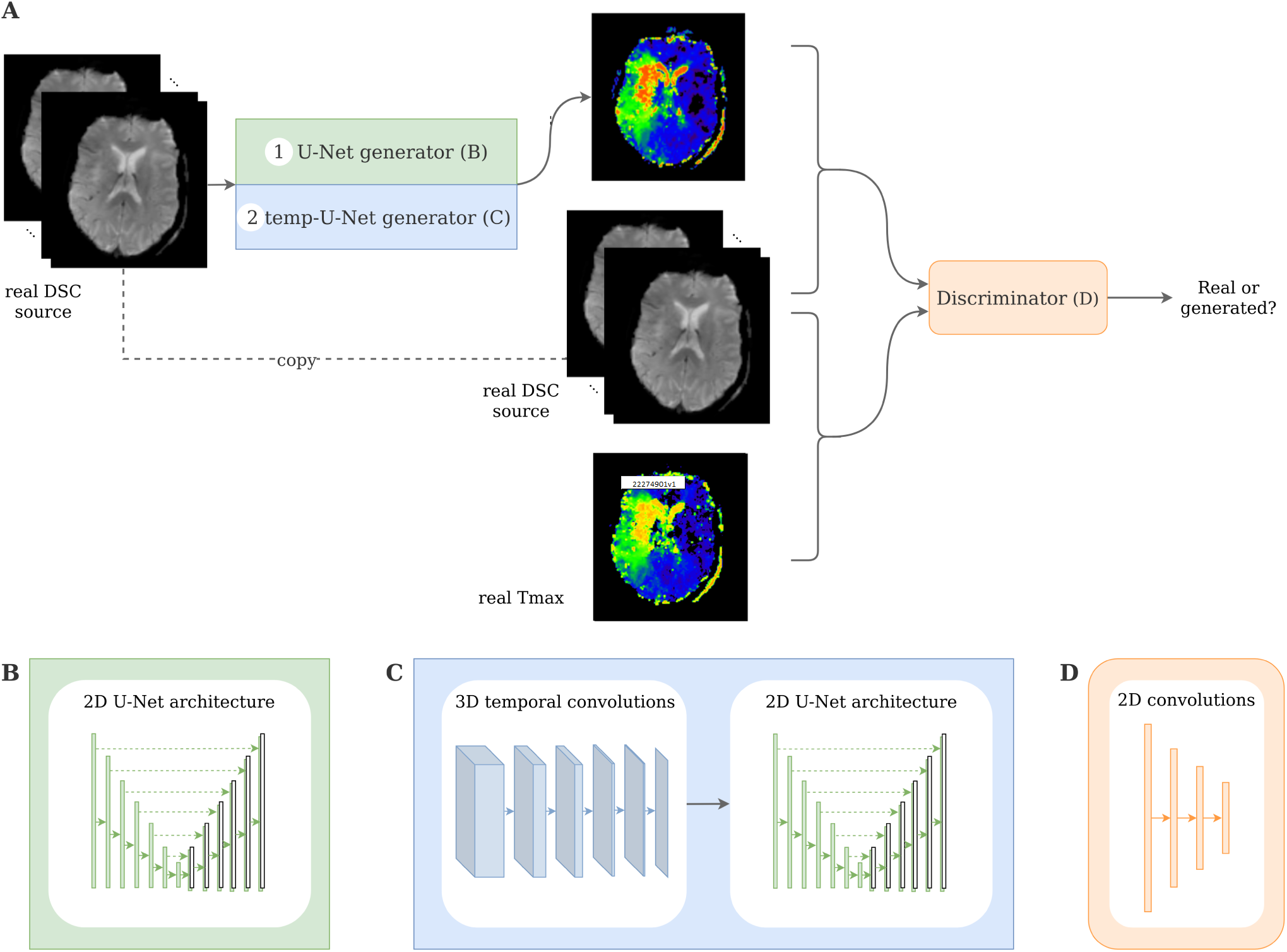
Architecture of the pix2pix and temp-pix2pix GAN. A shows the overall GAN architecture whereas B and C depict the two different generators and D the discriminator.

### 2.3 Network architecture

The GAN architecture was adapted from the pix2pix GAN (Isola et al. (2018)). In our first architecture we utilized the original U-Net generator as proposed in the paper with the time steps being represented in the channels. For the second architecture we modified the U-Net by adding 3D temporal convolutions before feeding the result into the U-Net in the generator (see Figure 2).

Both GAN architectures consisted of two neural networks: the generator G and the discriminator D. On the one hand, the generator’s task was to synthesize perfusion parameter maps such as Tmax or CBF from the DSC source image. The discriminator, on the other hand, learned to distinguish between the real DSC source image together with the real perfusion parameter map and the real DSC with the generated perfusion parameter map.

In general, the objective function of a conditional GAN such as the pix2pix GAN is:

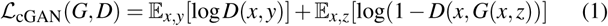

where *x* is the input image (DSC source in our case) and *y* the output image (Tmax for example) and *z* a noise vector. The generator tries to maximize the objective which is achieved when the discriminator outputs a high probability of the generated image pair being real and a low probability for the real image pair respectively. In contrast to that, the discriminator tries to minimize this objective and identify the real input images. The pix2pix GAN does not directly incorporate the noise vector *z* but introduces noise in the network using dropout in the generator.

The loss of the generator consisted of two parts. The first part was the adversarial loss which took into account the feedback of the discriminator as described above. Additionally, a reconstruction loss directly penalized deviation from the original image:

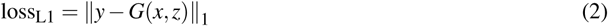

This second loss was added to the adversarial loss and weighted by a scalar *λ* which was set to 1. The pix2pix generator was a U-Net with 6 down- and upsampling layers (see Figure 2B). One DSC source slice at a time was fed as an input to the generator. The different time points of the DSC were concatenated in the channel dimension. Each downsampling layer consisted of a batch normalization layer as well as a LeakyReLU with slope 0.2 and the upsampling layers of ConvTranspose-layers, batch normalization and a ReLU activation. After the last convolution, a tanh was applied.

In contrast to that, the generator of the temp-pix2pix GAN took one slice of the DSC source at all time points as an input. The time sequence of slices was then fed through 6 3D convolutions over the time dimension iteratively reducing this dimension to 1. Each convolutional layer was followed by a batch normalization layer and a LeakyReLu with slope 0.2. After the temporal path, the output was fed into a 2D U-Net with convolutions over the spatial dimensions with 6 down- and upsampling layers as described above (see Figure 2C).

The discriminator adapted the architecture of the discriminator from the PatchGAN as suggested by Isola et al. (2018). It consisted of 3 convolutional layers with batch normalization and a LeakyReLU activation function followed by another convolutional layer and a sigmoid activation function (see Figure 2D). For both the generator and discriminator the kernel size was 4 with strides of 2.

### 2.4 Training

For each architecture, 5 GANs were trained on the acute stroke dataset from Heidelberg for each of the five parameter maps (CBF, CBV, MTT, Tmax and TTP). The models were trained for 100 epochs with a learning rate of 0.0001 for both generator and discriminator using the Adam optimizer with *β*_1_ = 0.5 and *β*_2_ = 0.999. The batch size was 4 and dropout 0. As the PEGASUS dataset was smaller, the models trained on the acute stroke data served as a weight initialization for the PEGASUS models and were then further trained for 50 epochs. Thus, in total, 10 models were trained per architecture.

All hyperparameters mentioned above were tuned and selected according to visual inspection and the performance on the validation set. Due to the computational limitations an automated search was not feasible. The code was implemented in PyTorch and is publicly available^2^. The models were trained on a TESLA V100 GPU (NVIDIA Corporation, Santa Clara, CA, USA).

### 2.5 Performance evaluation

The generated images were first visually inspected. Additionally, four metrics were applied: the mean absolute error (MAE) or L1 norm of the error, the normalized root mean squared error (NRMSE), the structural similarity index measure (SSIM) and the peak-signal-to-noise-ratio (PSNR).

The MAE is defined voxel-wise and measure the average absolute of the error between the real image *y* and the generated image *ŷ*:

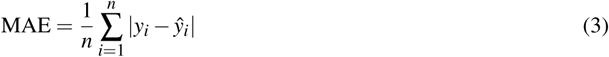

The NRMSE is defined as the root mean squared error normalized by average euclidean norm of the true image *y*:

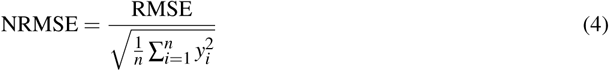

With

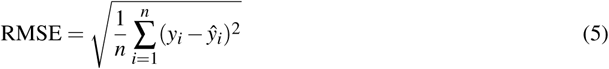

The SSIM is defined as a combination of luminance, contrast and structure and can be summed up as:

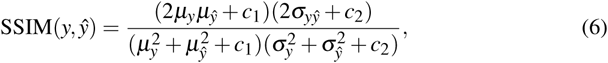

where *µ*_*y*_ and *µ*_*ŷ*_ are the average values of *y* and *ŷ* respectively, *σy* the variance and *σ*_*yŷ*_ the covariance. *c*1 and *c*2 are constants for stabilization and defined as *c*1 = (*k*1*L*)2 and *c*2 = (*k*2*L*)2 with *L* being the dynamic range of the pixel values and *k*_1_, *k*_2_ ≪1 small constants. The higher the SSIM, the more similar are the two images with 1 denoting the highest similarity. The PSNR is defined as:

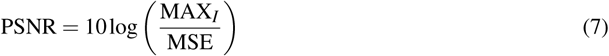

with MAX*I* being the maximal possible pixel/voxel value. It describes the ratio between the maximal possible signal power and noise power contained in the sample.

## 3 RESULTS

Visual inspection of the results of the acute stroke dataset showed that the perfusion parameter maps generated by the temp-pix2pix GAN looked similar to the ground truth (see Figure 3). For the pix2pix model, on the other hand, only the CBF, CBV and MTT were of sufficient quality, whereas the time-dependent parameters TTP and Tmax did not consistently resemble the ground truth (also Figure 3).

**Figure 3.**
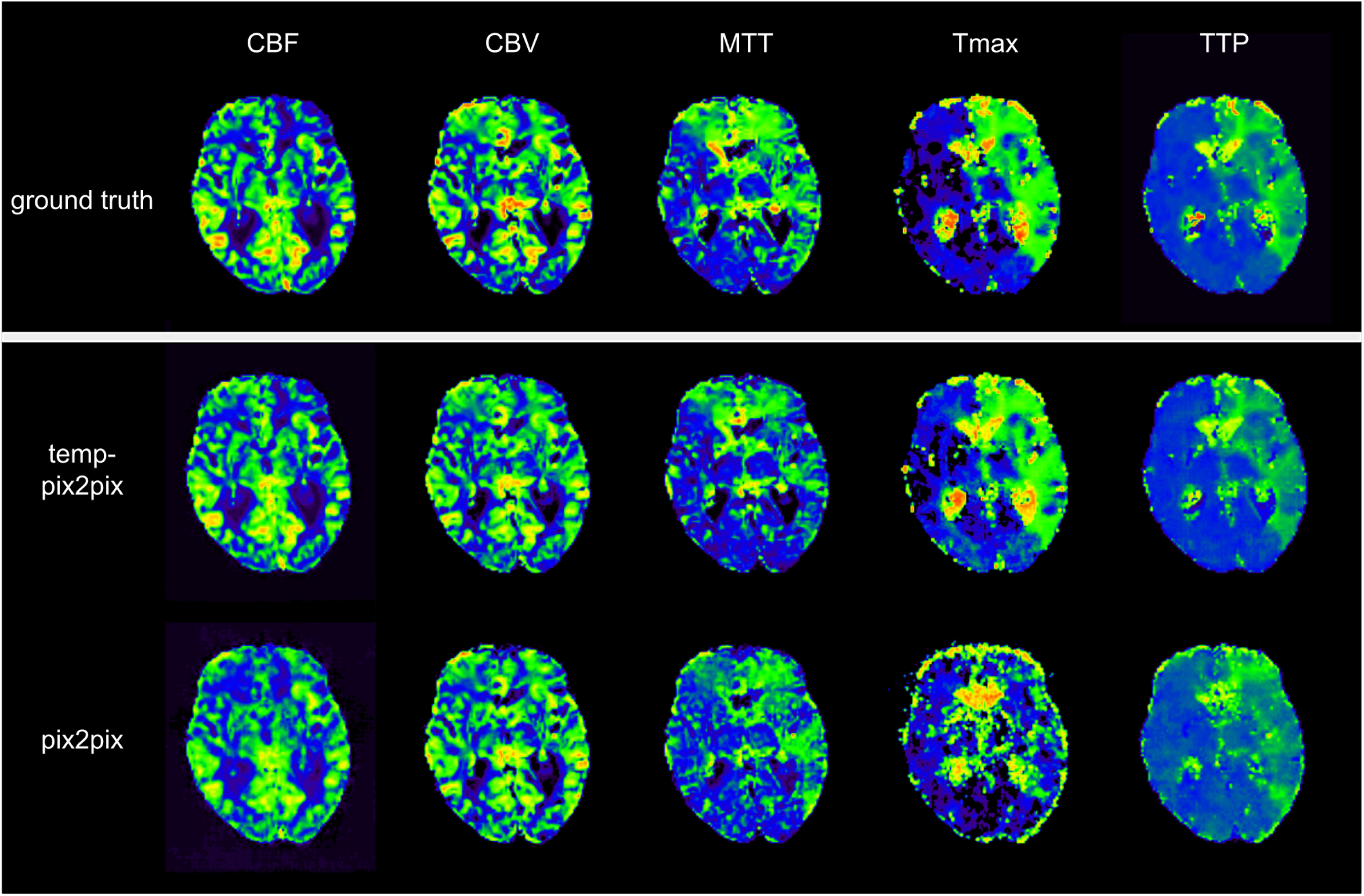
Synthesized perfusion parameter maps (middle and bottom row) compared to the ground truth reviewed by an expert (top row) for one representative patient from the acute stroke test dataset. The perfusion parameter maps generated by the temp-pix2pix all look similar to the ground truth whereas the time-dependent parameters (Tmax and TTP) are not well captured by the pix2pix GAN.

The quantitative analysis in the acute stroke dataset revealed for all parameter maps a high SSIM ranging from 0.92-0.99 for the temp-pix2pix model (Figure 4). In contrast to this, the pix2pix GAN showed a comparable or worse SSIM ranging from 0.86-0.98. A performance difference between the pix2pix and temp-pix2pix model was especially prominent for Tmax and TTP (SSIM 0.92 vs 0.86 and 0.95 vs 0.91, respectively). For the PEGASUS dataset, the perfusion maps generated by both the fine-tuned pix2pix and temp-pix2pix GAN look similar to the ground truth (see Figure 5). For both networks, MTT appeared to be the least well reconstructed parameter map which is also reflected in the metrics (Figure 6). Furthermore, the high intensities of Tmax were not well captured by the pix2pix GAN (Figure 5). The performance metrics of the pix2pix and temp-pix2pix GAN and the ground truth for the PEGASUS dataset showed low error and high SSIM and PSNR for CBF, CBV and Tmax. Here, for most metrics, the temp-pix2pix GAN achieved a slightly better performance in contrast to the pix2pix GAN. For MTT and TTP the temp-pix2pix showed a better performance compared to the pix2pix GAN (SSIM 0.84 vs 0.78 and 0.86 vs 0.82 respectively). Overall, the metrics of the synthesized MTT and TTP maps obtained a worse performance compared to the other parameter maps. Figure 7A showed two patients whose generated parameters showed the worst performance. For the acute stroke dataset these are two Tmax maps (Figure 7A, first and second column). Whereas the generated Tmax in the first column did not capture the high intensities well, the generated map in the second column visually looked well. For the PEGASUS models, MTT performed the worst (Figure 7A, third and fourth column). In the third column the generated MTT appears less noisy than the ground truth. In contrast to that, in the fourth column the generated MTT map looked noisier compared to the ground truth. Figure 7B showed the Tmax maps generated by the temp-pix2pix and pix2pix GAN for four patients for which an AIF could not be placed.

**Figure 4.**
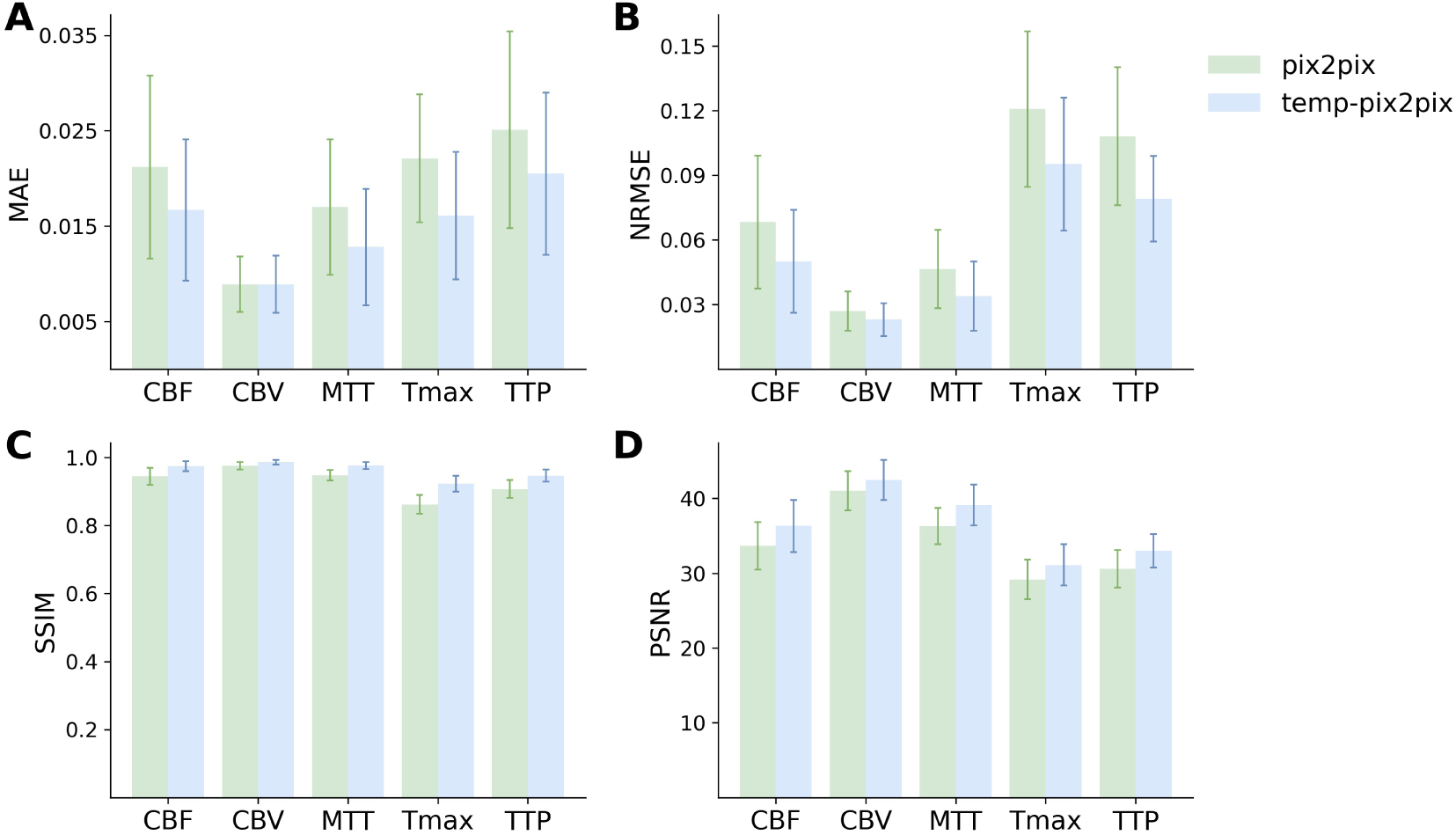
Mean performance metrics for evaluating the similarity between the ground truth and the synthesized parameter maps generated by the pix2pix GAN (green) and the temp-pxi2pix GAN (blue) on the acute stroke dataset. A and B show the mean absolute error (MAE) and normalized mean root squared error (NRMSE) respectively (the lower the better). C and D show the structural similarity index measure (SSIM) and the peak-signal-to-noise-ratio (PSNR) (the higher the better). For all parameter maps the temp-pix2pix architecture shows a better or comparable performance compared to the pix2pix GAN. For the time-dependent parameter maps Tmax and TTP the difference between the pix2pix and temp-pix2pix GAN performance is larger than for the other three maps. The errorbar represents the standard deviation.

**Figure 5.**
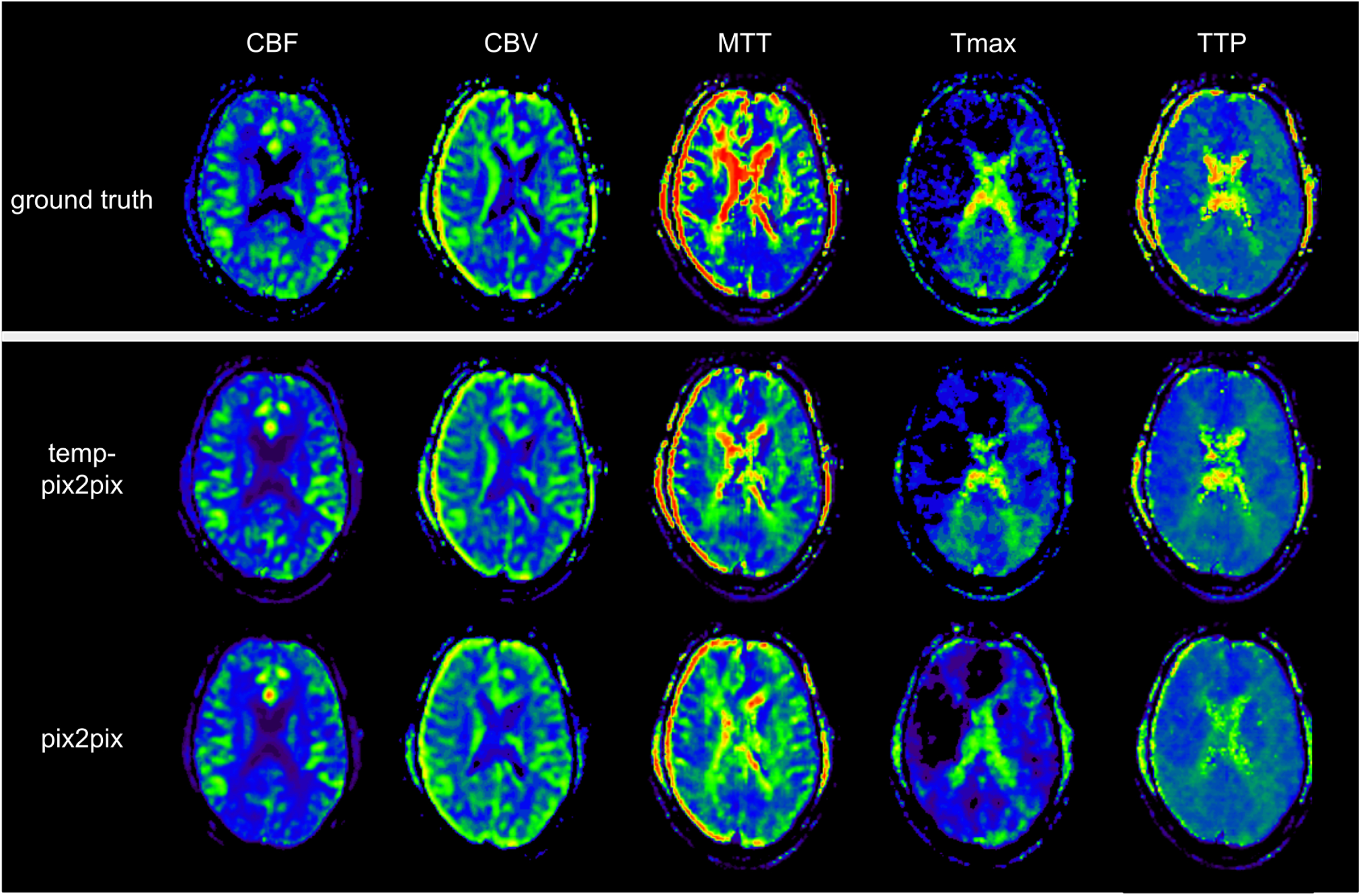
Synthesized perfusion parameter maps (middle and bottom row) compared to the ground truth reviewed by an expert (top row) for one representative patient from the PEGASUS test dataset. Both pix2pix and temp-pix2pix GAN synthesized most parameter maps that resemble the ground truth. Parts of MTT were not entirely captured by pix2pix and temp-pix2pix. Moreover, the pix2pix GAN did not synthesize the higher intensities of Tmax well. For MTT and Tmax, the temp-pix2pix GAN showed better performance in all metrics compared to the pix2pix GAN.

**Figure 6.**
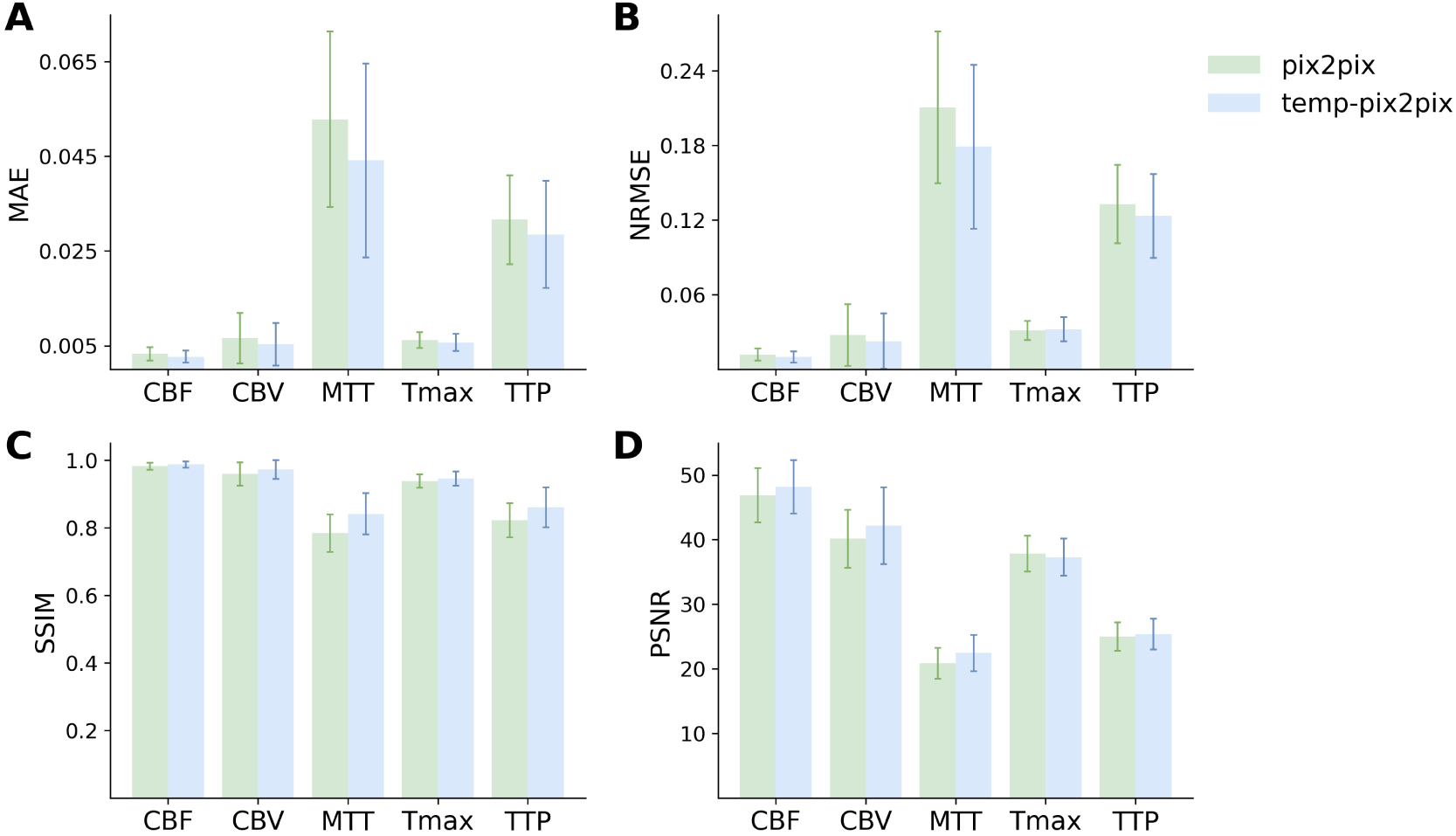
Mean performance metrics for evaluating the similarity between the ground truth and the synthesized parameter maps generated by the pix2pix GAN (green) and the temp-pxi2pix GAN (blue) on the PEGASUS dataset. A and B show the mean absolute error (MAE) and normalized mean root squared error (NRMSE) respectively (the lower the better). C and D show the structural similarity index measure (SSIM) and the peak-signal-to-noise-ratio (PSNR) (the higher the better). For most metrics and parameter maps the temp-pix2pix architecture shows a better performance compared to the pix2pix GAN. In terms of the metrics, the generated MTT maps showed the worst performance. The errorbar represents the standard deviation.

**Figure 7.**
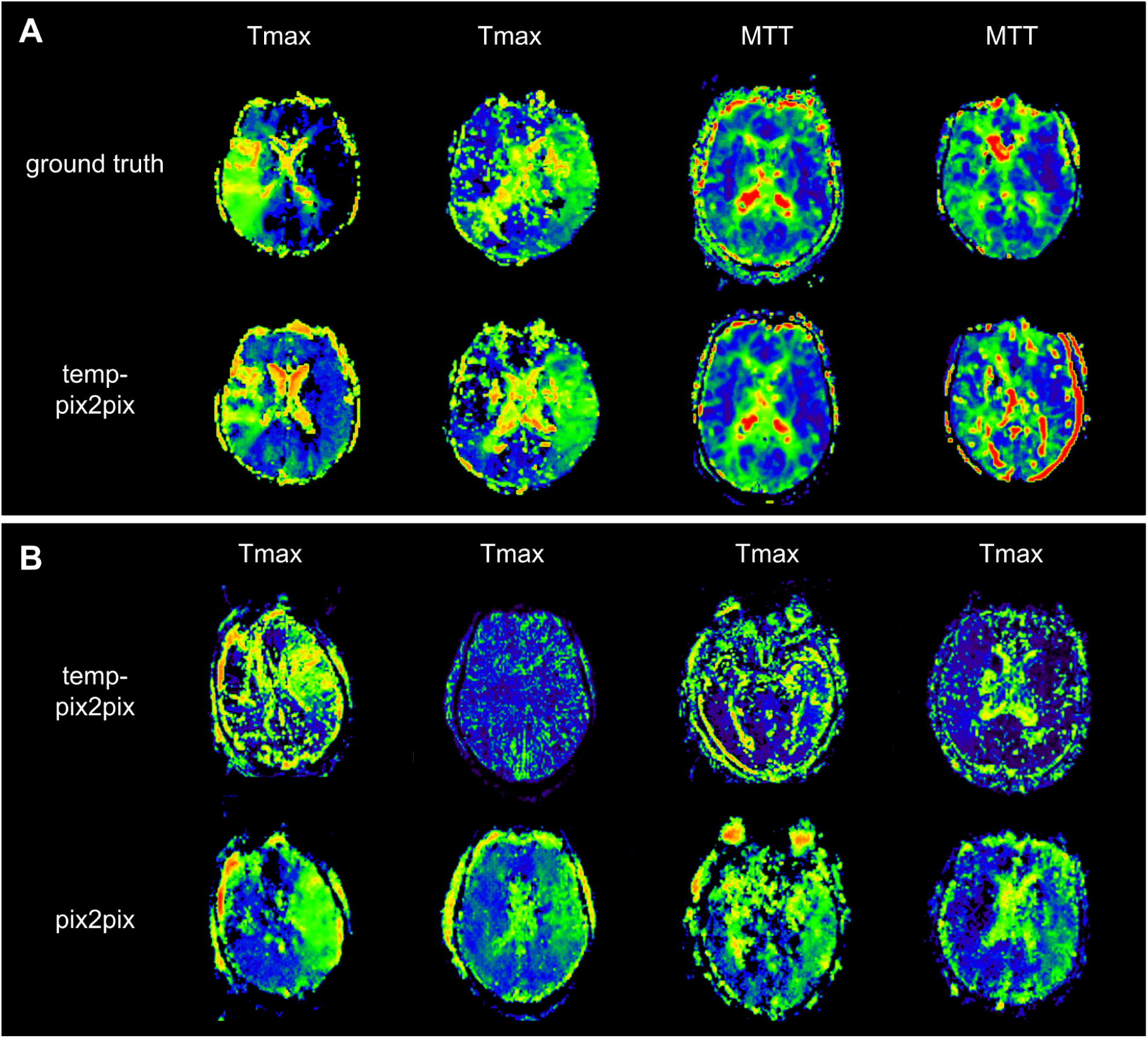
The two patients with the poorest performance according to the metrics for each of the two datasets (A) and patients for which no AIF could be computed (B). A: The first and second column show Tmax for two acute stroke patients. Whereas the synthesized image in the first column does not fully capture the hypoperfused areas, the generated image in the second column looks quite close to the ground truth. Column three and four show MTT for two PEGASUS patients. While the generated image in the third column shows less noise than the ground truth, the GAN introduced noise in the fourth column in the synthesized image. B: Four Tmax maps generated by temp-pix2pix (upper row) and pix2pix (lower row) for cases from the acute stroke data for which no AIF could be computed and, thus, with conventional methods not imaging would be available. Note that since motion artifacts affect the quality of the time-series, in these cases the baseline pix2pix performs better than the temp-pix2pix.

## 4 DISCUSSION

In the present study, we propose a novel pix2pix GAN variant with temporal convolutions - coined temp-pix2pix - to generate expert-level perfusion parameter maps from DSC-MR images in an end-to-end fashion for the first time. The temp-pix2pix architecture showed high performance in a dataset of acute stroke patients and good performance on data of patients with chronic steno-occlusive disease. Our results mark a decisive step towards the automated generation of expert-level DSC perfusion maps for acute stroke and their application in the clinical setting.

In acute stroke, “time is brain” (Saver (2006)). This requires rapid decision making in the clinical setting to ensure an optimal outcome for an affected patient. In such a situation, when DSC perfusion-weighted imaging is used to stratify patients for treatment, a major bottleneck is the generation of parameter maps derived from the DSC source images. These maps of TTP, CBF, CBV, MTT, and Tmax are different representations of the information encoded in the time-intensity curve for each voxel. For all except TTP, to derive robust and valid parameter maps, the time-intensity curve must be deconvolved with an AIF (Calamante (2013)). Ideally, the AIF is derived for each voxel separately, but in the clinical setting the calculation of a global AIF is preferred (Calamante (2013)). The gold standard is the manual selection of several - usually 3 or 4 - AIFs in the hemisphere contralateral to the stroke, from segments of the middle cerebral artery (Calamante (2013)). The manual selection of AIFs is a tedious and time-consuming process that can only be performed after training (Calamante (2013)). Therefore, automated methods whose results are subsequently reviewed by an expert are preferred in clinical practice (Calamante (2013)). While automated methods have shown inconclusive results in the literature (Hansen et al. (2016); Ghodrati et al. (2021); Galinovic et al. (2012); Pistocchi et al. (2022); Deutschmann et al. (2021)), they are successfully used in acute stroke to identify stroke-affected tissue. In our sample, this was confirmed as the AIFs only required expert adjustment in two out of 204 patients in the acute stroke set. Nevertheless, this approach still requires a manual check resulting in a time delay of a few minutes per patient before patient stratification. As a consequence, there is a major clinical need for automated methods that provide final perfusion parameter maps without any manual input. Here, we chose a GAN AI approach, as presenting this methodology expert level perfusion maps would lead to a model after training that could then *generate* expert-level perfusion maps, implicitly encoding the choice of AIFs within ∼1.8 seconds per patient. Our exploratory results show that this approach was successful.

This may have a positive impact on the clinical setting. First, it would eliminate the need for manual review of AIFs. This would reduce the time needed to calculate perfusion parameter maps and also reduce resource requirements as radiologists and neurologists would no longer need to be trained on how to identify optimal AIFs. Second, as we have shown, it is even possible to calculate parameter maps for patients who currently have to be excluded due to motion artifacts that make it impossible for the standard software to calculate the parameter maps. At this point, it is important to emphasize that our study is exploratory and the generated model was and is only used for internal research purposes. This is due to the fact that the generative AI has fundamentally learned to approximate the non-AI algorithm that was originally used to calculate the perfusion parameter maps. To maximize clinical impact, we thus encourage the developers and vendors of relevant clinically used perfusion software to consider adding GAN-based automated perfusion calculation modules to their products. To facilitate this process, we have made our code publicly available.

One of the most important contributions of our approach was the consideration of the temporal dimension of the time series input. Not surprisingly, the temp-pix2pix architecture performed better than the pix2pix GAN without a temporal component in both datasets. This was particularly noticeable in the acute stroke dataset for parameters directly related to the correct order of the time intensity curve, namely TTP and Tmax. Maps of CBF, CBV and MTT (derived by the central volume theorem as CBV/CBF) also performed quite well in the baseline architecture without a temporal component, as for these maps the order of input is not as relevant. This is because CBV corresponds to the area under the time intensity curve and CBF is calculated based on the height of the slope, which are indifferent to the order. In the chronic stroke dataset, the temp-pix2pix also outperformed the baseline GAN without a temporal component. However, the difference in performance was not as pronounced as in the acute stroke dataset. This could be due to the fact that patients with acute vascular obstruction usually have significantly higher delays than patients with chronic steno-occlusive disease, and the performance advantage of temp-pix2pix increases with increasing delay. It is noteworthy that in contrast to the acute stroke patients in the chronic steno-occlusive cohort, MTT and TTP maps performed worse than the other parameter maps. This might be related to the more complex perfusion pathophysiology in chronic steno-occlusive disease. Whereas in acute stroke, delay is the main contributor to blood flow abnormalities, in chronic steno-occlusive disease it is the sum of delay and considerable dispersion due to vessel abnormalities (Calamante et al. (2006)). This could pose particular difficulties for neural networks to learn the relationships required to create parameter maps: MTT is as a parameter that depends on two other parameters (CBV and CBF) in the original software solutions, which are likely to have greater variability in chronic steno-occlusive disease. Addtionally, TTP delays are attributable to both delay and dispersion, with varying weights in individual patients leading again to a larger variability (this effect is much less pronounced in Tmax parameter maps due to the deconvolution procedure). Such increased variability might lead to less stable models and thus increased noise in the generated maps.

Our work is the first work to utilize GANs to create perfusion parameter maps in DSC-imaging. A few works exist that used different machine learning and deep learning methods to generate parameter perfusion maps from the DSC source image. For instance, McKinley et al. (2018) used several classical voxel-wise machine learning approaches to generate manually validated perfusion parameter maps and identified a tree-based algorithm as the best performing model. Their best results for Tmax achieved a lower performance with a NRMSE of 0.113 compared to our best model with a NRMSE of 0.095. Vialard et al. (2021) suggested a deep learning based spatiotemporal U-net approach for translating DSC-MR patches to CBV maps in patients with brain tumors. With a SSIM of 0.821 their generated CBV maps obtained a worse performance compared to our CBV generated by the temp-pix2pix model with a SSIM of 0.986. In the field of stroke, Ho et al. (2016) proposed a patch-based deep learning approach to generate CBF, CBV, MTT and Tmax. The average RMSE for their generated Tmax showed a higher error of 1.33 compared to ours with 0.06. Hess et al. (2019) utilized a different voxel-wise deep learning approach to approximate Tmax from DSC-MR. This approach was clinically evaluated in another study (Meier et al. (2019)). In Hess et al. (2019) they reported the performance in terms of MAE with clipping to not account for noise. Their generated Tmax achieved a MAE with clipping of 0.524 compared to our approach showing a MSE of 0.016. These differences compared to our study might be due to the novel use of the GAN method and the fact that our model considered whole slices instead of patches to better account for the spatial dimension. Our study has several limitations. First, our network was based on 2D slices instead of the full 3D volumes due to computations restrictions. It is likely that results could be improved further using the full 3D images. Secondly, our study is an exploratory hypothesis generating study. Its results need to be clinically validated in a future study before an integrating into clinical practice would be possible. Lastly, our approach so far is a black-box approach. It could be extended with explainable AI to generate insights which areas in the source images are particularly relevant for the creation of different perfusion parameter maps. This could further elucidate the causes of the performance differences between maps that we identified and could guide the way for further improvements.

## 5 CONCLUSION

We generated expert-level perfusion parameter maps using a novel GAN approach showcasing that AI approaches might have the ability to overcome the need for oversight by medical experts. Our exploratory study paves the way for fully-automated DSC-MR processing for faster patient stratification in acute stroke. In the clinical setting where time is crucial for patient outcome, this could have a big impact on standardized patient care in acute stroke.

## Data Availability

All data produced in the present study are available upon reasonable request to the authors

## ACKNOWLEDGEMENTS

Computation has been performed on the HPC for the Research cluster of the Berlin Institute of Health.

## Footnotes

2 https://github.com/prediction2020/DSC-to-perfusion

